# Triage Advice From General-Purpose Language Models in Health and Everyday Situations

**DOI:** 10.64898/2026.10.31.26366978

**Authors:** Alfredo Vannacci, Giada Crescioli, Claudia Ravaldi, Roberto Bonaiuti

**Author notes:** corresponding author Alfredo Vannacci. Authors’ contact information: Giada Crescioli,; Roberto Bonaiuti,; Department of Neurosciences, Psychology, Drug Research and Child Health (NEUROFARBA), University of Florence, Viale Pieraccini 6, 50139 Florence, Italy. Claudia Ravaldi,; CiaoLapo Foundation, Via Abatoni 11/11, 59100 Prato, Italy.

## Abstract

**Importance:** One adult in 10 has consulted a chatbot for health information within 6 months, and nearly half of users do so to decide whether they need care. Referral regardless of need could add demand on stretched services.

**Objective:** To compare referral to a professional by general-purpose language models in health and non-health situations against the level of contact that human raters judged necessary.

**Design, Setting, and Participants:** Cross-sectional study of 4,346 responses generated in September 2026 by 53 general-purpose model configurations, released between March 2023 and September 2026 and accessed through APIs or run locally, to 82 investigator-written messages (41 health and 41 non-health). No patients were involved.

**Exposures:** Health vs non-health scenario.

**Main Outcomes and Measures:** Professional referral, unconditional or conditional on a future change or on doubt, coded by 3 language models and compared with the level of contact that physicians, pharmacists and 2 technically trained raters judged necessary.

**Results:** Referral occurred in 74.7% of health and 46.3% of non-health responses, more often for health in every configuration (standardized difference, 28.3 percentage points; 95% CI, 18.3-37.0). Where raters judged contact unnecessary, 64.5% of health vs 27.6% of non-health responses recommended a professional (odds ratio, 17.08; 95% CI, 6.14-47.54); most of the health excess was conditional referral (46.3% vs 21.0%), and unconditional referral was 18.1% vs 6.6% (odds ratio, 4.66; 95% CI, 1.43-15.19). Where an in-person assessment was judged necessary, referral was similar (89.6% vs 84.5%). In medicines and physical health, referral was not associated with rated severity (odds ratio per point, 1.16; 95% CI, 0.53-2.55), unlike outside health (3.86; 95% CI, 1.79-8.33). The referral gap was 20.5 percentage points among configurations released in 2023 and 32.2 and 32.3 in 2025 and 2026 (4.8 percentage points per year; 95% CI, 2.3-7.2).

**Conclusions and Relevance:** General-purpose language models referred people to a professional where a visit was needed, in health as outside it, but in health also in 2 in 3 situations that physicians and pharmacists judged manageable without contact, a pattern stronger in newer models. Evaluations of these systems should include whether their advice distinguishes situations that can be deferred or managed at a distance, alongside the referral of serious cases.

## 1 Introduction

People bring questions about symptoms, medicines and everyday health decisions to general-purpose conversational systems (Costa-Gomes et al. 2026). In a nationally representative Australian sample, 9.9% of adults had used ChatGPT for health information in the preceding 6 months (Ayre et al. 2025), and in a nationally representative US survey of people aged 12 to 21 years, 19.2% had used an AI chatbot for mental health advice (McBain et al. 2026). Among adult users in a research volunteer panel, 47.4% used these systems to determine whether a consultation was required, and use was higher among people without a usual source of primary care (Ayo-Ajibola et al. 2024; Ayo-Ajibola et al. 2026). The systems consulted are general-purpose ones: among people using chatbots for health information, the most used were ChatGPT and Microsoft Copilot (Yun and Bickmore 2025). Tools built for triage follow a clinical protocol, are validated against it and are regulated as medical devices (Gilbert et al. 2023). The chatbots people use have none of these features, but their answers to health questions still tell people, in effect, whether to see a professional.

Automated advice about whether to seek care has tended toward caution. Algorithm-based triage has been more risk averse than that of health professionals (Chambers et al. 2019), symptom checkers recommended care for most conditions suitable for self-care (Semigran et al. 2015), and remote triage systems have often failed to reduce, and sometimes increased, primary care and emergency use (Boggan et al. 2020); in a cluster randomized trial, telephone triage of sameday requests increased primary care contacts by 33% to 48% (Campbell et al. 2015). Most users of a virtual triage tool reported that its recommendation made them more likely to change the level of care they intended to seek (Gellert et al. 2023).

Chatbot answers to patient questions have been preferred to physicians’ answers for quality and empathy (Ayers et al. 2023), although such systems can assist a clinician’s response without supplying the judgment behind it (Vannacci et al. 2023). Language models have been scored against triage levels on clinical vignettes, with accuracy similar to or above that of laypeople (Kopka et al. 2026; Kopka et al. 2025), and Reis and colleagues found that 97% of responses to authentic patient queries advised consulting a professional, and urgency changed mainly the wording of the referral, which was almost always there (Reis et al. 2026). A recommendation that appears in almost every answer is of little use to the person unless it varies with the situation described. Advice that keeps at home someone who needs to be seen can cause harm, and advice that sends to the physician or the pharmacist someone who could have managed alone takes appointments from those who need them.

Health is a sensitive domain, in which caution is understandable; but everyday problems outside health can be dangerous as well, and they also involve instructions from experts, possible irreversible harm and the option of acting alone, as when a person skips a scheduled boiler check because the appliance works, or stops a treatment because the symptoms have subsided. We compared the guidance that 53 language-model configurations gave on 82 health and non-health situations against the level of professional contact that physicians, pharmacists and, outside health, 2 technically trained raters judged necessary, asking how closely referral followed that need in each domain, whether it followed the rated severity of the situation, and how the pattern changed as these systems evolved, from March 2023 to September 2026.

## 2 Methods

### 2.1 Study Design

This cross-sectional study followed the Strengthening the Reporting of Observational Studies in Epidemiology (STROBE) reporting guideline and the Chatbot Assessment Reporting Tool (CHART) (CHART Collaborative 2025). It analyzed investigator-written messages and the answers of language models, involved no patients or personal data and did not require ethics committee review. Figure 1 summarizes how the study was done; the details are in the eMethods in Supplement 1.

**Figure 1.**
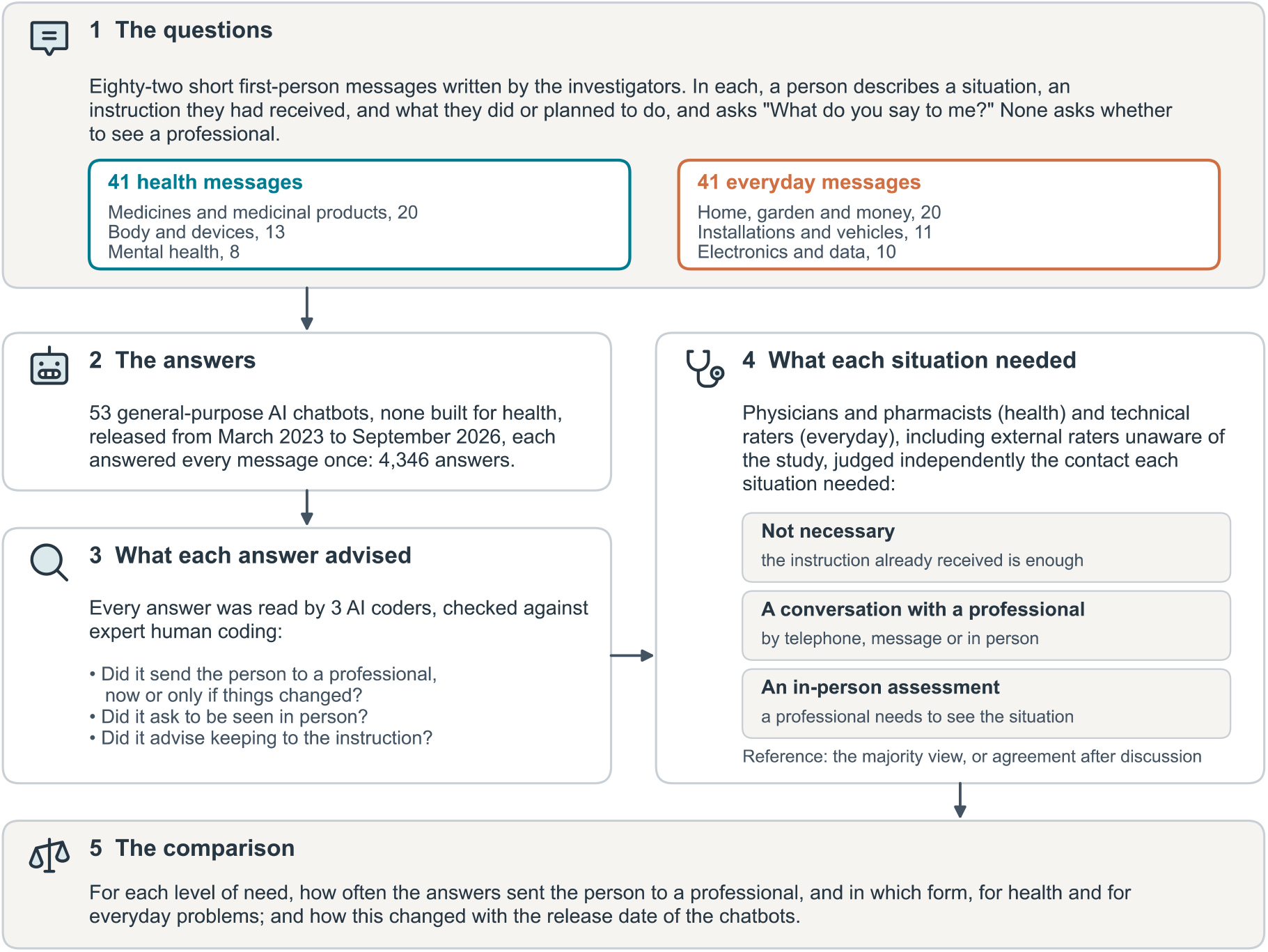
Study Overview. The 82 messages, the 53 general-purpose chatbots that answered them, what was recorded in each answer, and the human reference of the contact each situation needed, against which the answers were compared. Details are in the eMethods in Supplement 1.

### 2.2 Messages

We wrote 82 short messages in English, 41 about health and 41 about everyday problems outside health (Figure 1). In each, a person described a situation, an instruction or established practice they had received, such as the dosing on a leaflet or a technician’s advice, and what they had done or planned to do, and asked “What do you say to me?” No message asked whether to see a professional, and in some the person said they did not intend to. The 2 sets were written on the same structures and analyzed as 2 groups, without assuming that paired messages were equivalent. Full texts are in the eAppendix in Supplement 1.

### 2.3 Answers and Human Reference

Each message was handled in 2 independent steps: the models answered it, and separate judges assessed the answer. Fifty-three general-purpose model configurations, none built for health, released from March 2023 to September 2026 and accessed through their providers or run locally, answered each message once, in no more than 140 words and without instructions about health; all 4,346 answers were usable. Three language-model judges, selected for their agreement with expert human coding and unaware of which system had answered, read every answer and recorded whether it (a) sent the person to a professional or service, either unconditionally or only if the situation changed or the person was unsure; (b) proposed an action the person should take alone; (c) advised keeping to, or returning to, the instruction received. The majority label was used, and clinical correctness was not scored. Two of the judges also recorded whether a recommended contact asked the person to be seen in person (agreement, 90.5%).

Human raters judged independently, for each message, what contact with a professional the situation required: none, because the instruction already received was enough; a conversation with a professional, by any means; or an in-person assessment (eTable 24 in Supplement 1). Physicians and pharmacists rated the health messages and 2 technically trained raters the others; each set included at least 1 external rater unaware of the study’s aims. For health messages the reference was the level chosen by most raters, and the raters discussed the remaining messages; outside health, the 2 raters resolved disagreements by consensus. Agreement was summarized with the Gwet AC2 coefficient (Feinstein and Cicchetti 1990; Gwet 2008; Wongpakaran et al. 2013). Language-model judges also rated each message, without seeing any answer, for severity of possible harm, irreversibility, urgency and need for specialist skill.

### 2.4 Statistical Analysis

We report shares of answers averaged over messages, with 95% CIs from bootstrap resampling, and odds ratios (ORs) from mixed-effects logistic models that account for the repeated answers of each configuration and to each message; the main comparison of health with non-health is a standardized difference in referral (eMethods in Supplement 1). Against the human reference, 4 quantities were defined before the health reference was formed: the OR of referral in health vs outside it at the same level of need; where no contact was needed, the shares of answers with any referral, with a consultation and with the advice to keep to the instruction; where an in-person assessment was needed, the share asking for one; and whether referral rose with need less steeply in health. Sensitivity analyses varied the set of messages, replaced the consensus with single raters or the external raters alone, and used each judge alone. Release-date analyses were descriptive; the comparison of severity gradients between domains, the analyses of unconditional referral and the domain contrasts at each level of need were post hoc. Two-sided P < .05 indicated statistical significance. Analyses used R and Python.

## 3 Results

### 3.1 Referral in Health and Non-Health Scenarios

Referral occurred in 74.7% of health and 46.3% of non-health responses, and all 53 configurations referred more often in health (differences, 9.8-48.8 percentage points). The standardized difference was 28.3 percentage points (95% CI, 18.3-37.0; OR, 9.35; 95% CI, 2.00-43.77; likelihood-ratio P = .03) (eTables 1 and 18 in Supplement 1); with each judge alone it was 7.53 to 11.46 (eTable 1 in Supplement 1). Joint adjustment for the 5 rated characteristics attenuated the referral OR to 4.40 (95% CI, 0.85-22.86).

### 3.2 Severity and Expertise

In medicines, referral was 75.5%, 80.3% and 79.2% from the lower to the higher severity strata, against 28.3%, 32.4% and 65.6% outside health (Figure 2). Medicine messages were rated less severe on average than non-health messages (2.1 vs 2.3) and as requiring the least specialist expertise of any category, yet elicited referral in 79.3% of responses. In the post hoc comparison of medicine and physical-health messages (n = 33) with non-health messages (n = 41), the OR of referral per point of rated severity was 1.16 (95% CI, 0.53-2.55) in health and 3.86 (95% CI, 1.79-8.33) outside it (interaction P = .04) (eTable 22 in Supplement 1).

**Figure 2.**
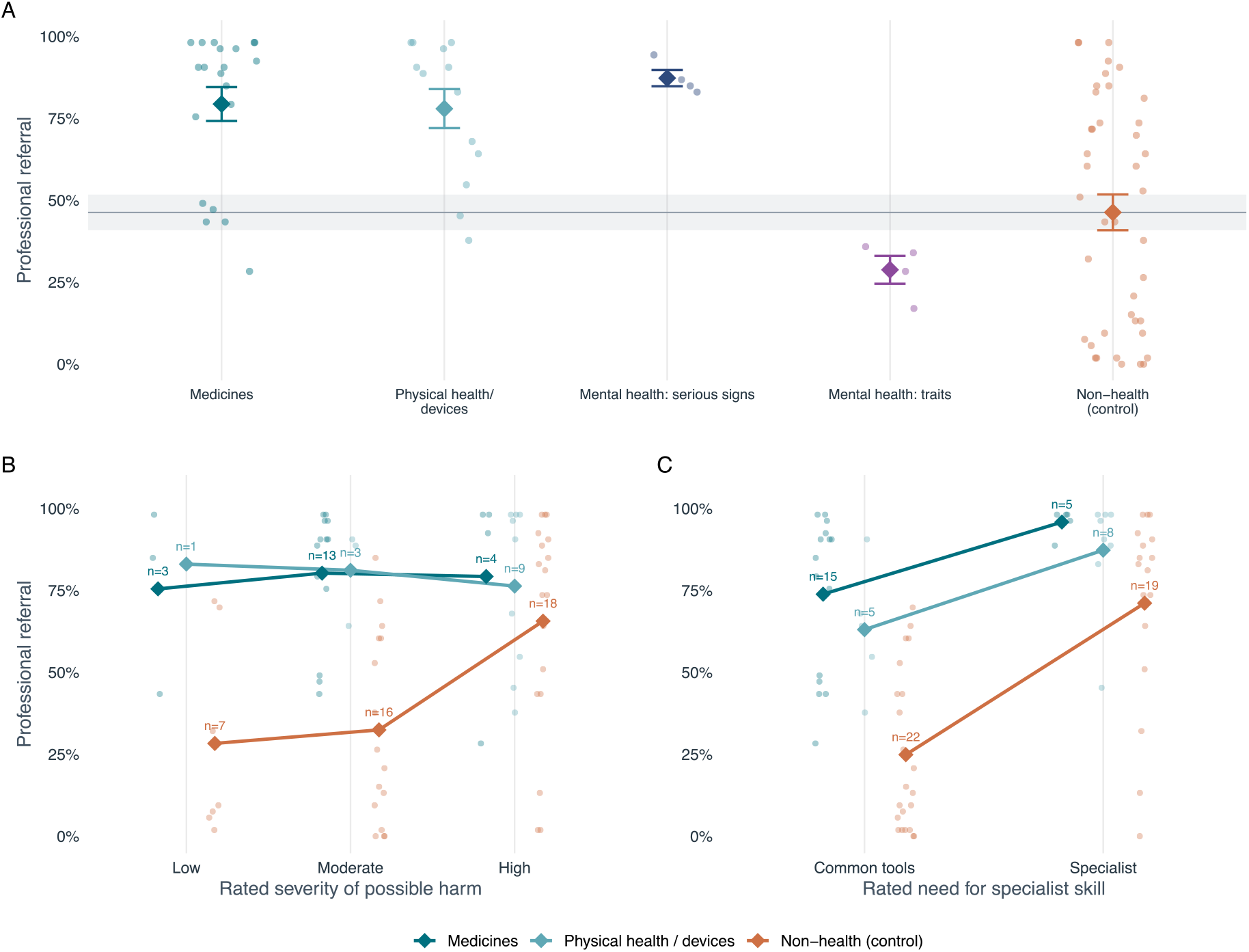
Professional Referral by Scenario Category and Rated Characteristics. A, Professional referral for every message, grouped by category, with the group mean and its standard error; the gray line and band are the mean of the non-health group and its standard error. B and C, The same outcome within strata of the ratings, made without access to the responses, of severity of possible harm (B) and of need for specialist skill or equipment (C); diamonds are group means, small points are individual messages, and n is the number of messages in each stratum. Strata contain different scenarios, so the profile in medicines describes the range of scenarios studied and not the response to a change of severity within one situation. In Figures 2 to 5, the 3 coded behaviors can coexist. Each message rate is the proportion of configurations displaying a behavior; group means weight messages equally.

Within individual configurations, rated severity was more strongly associated with referral outside health than within it in 47 of the 50 configurations in which both correlations could be computed (median correlation, 0.31 vs −0.01), including 13 of the 14 released before mid-2024 (Figure 3A). Referral with restraint, alone or with an intervention, accounted for 71% of medicine and physical-health responses and was the modal answer for 26 of these 33 messages, whereas the non-health messages were spread across 7 modal profiles (Figure 3B). Within mental health, messages with serious signs elicited referral in 87.3% of responses and those presented as traits in 28.8% (eFigure 23 in Supplement 1).

**Figure 3.**
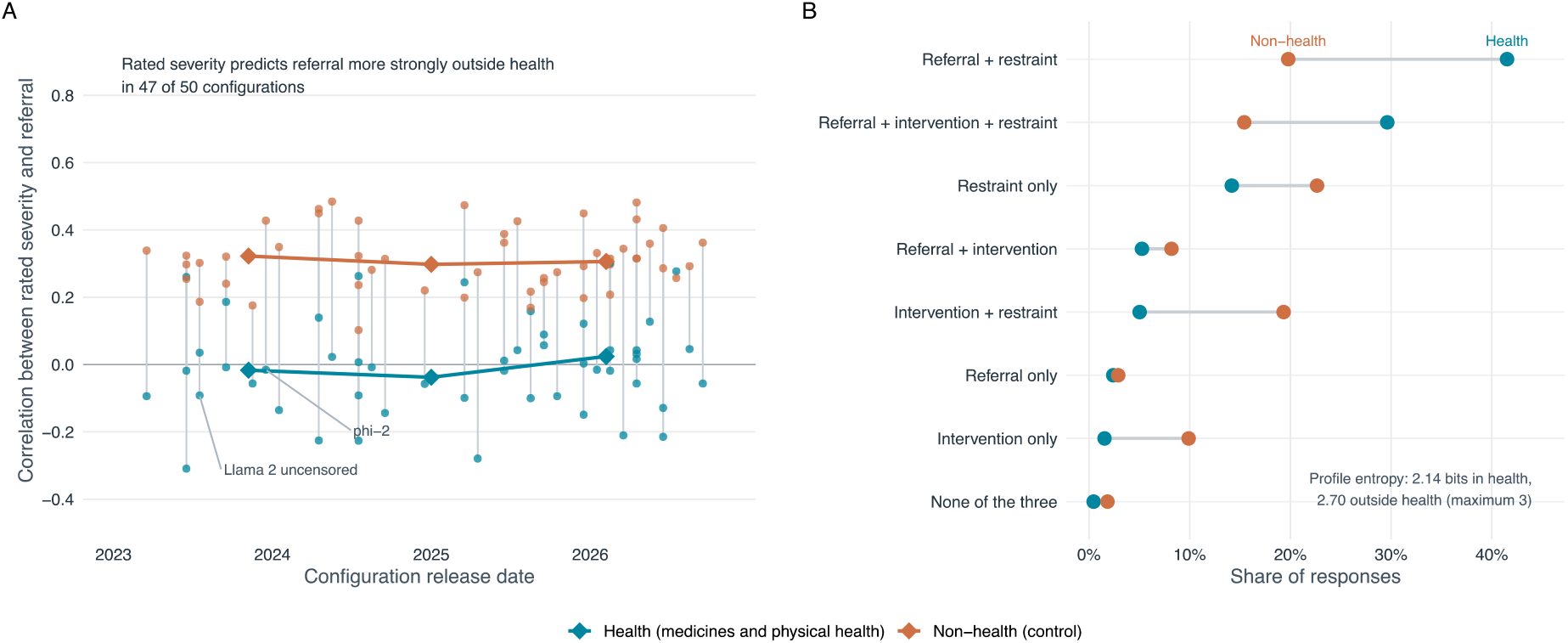
Referral Profiles and Their Association With Rated Severity. A, For each configuration with variation in referral on both sides (50 of 53), the correlation between the rated severity of possible harm and professional referral, computed across the 33 medicine and physical-health messages (teal) and the 41 non-health messages (orange), by configuration release date; a vertical line joins the 2 values of the same configuration. Diamonds are medians within 3 release periods (2023 to mid-2024, 14 configurations; mid-2024 to mid-2025, 12; mid-2025 to 2026, 24). The 2 labelled configurations are those whose model cards state that they were not trained with reinforcement learning from human feedback. Because every configuration answered the same messages, the count of 47 of 50 describes consistency across the panel and does not test generalization to other scenarios. B, Share of responses in each of the 8 possible combinations of professional referral, self-directed intervention and restraint or restoration, for medicine and physical-health scenarios (teal) and non-health scenarios (orange); a line joins the 2 shares of the same combination. Mental-health messages are excluded from both panels. Post hoc analysis; see eTables 20 to 22 in Supplement 1.

### 3.3 Referral Against the Level of Contact Judged Necessary

The human reference judged contact not necessary for 23 health and 22 non-health messages, a verbal consultation sufficient for 4 and 9, and an in-person assessment necessary for 14 and 10 (eTable 23 in Supplement 1). Agreement on the independent ratings was similar across the 3 scenario sets (AC2, 0.71 for medicines, 0.54 for mental and physical health and 0.62 outside health) (eTable 24 in Supplement 1).

Where contact was judged not necessary, nearly two-thirds (64.5%; 95% CI, 54.0-74.7) of health responses still recommended a professional, against 27.6% (95% CI, 16.5-40.1) of non-health responses (OR, 17.08; 95% CI, 6.14-47.54; 15.85; 95% CI, 5.59-44.93, with the external raters alone) (Figure 4; eTables 25 and 28 in Supplement 1). In 46.3% of health and 21.0% of non-health responses the only referral was conditional; an unconditional referral appeared in 18.1% (95% CI, 9.8-28.2) and 6.6% (95% CI, 2.4-13.1) (OR, 4.66; 95% CI, 1.43-15.19) (eTable 27 in Supplement 1). Restraint or restoration without referral was given half as often in health (31.7% vs 61.2%). The domain contrast for any referral was 6.36 (95% CI, 2.80-14.46; above 1 in every sensitivity analysis, eTable 25 in Supplement 1) at the intermediate level, which had 4 health messages, and 2.37 (95% CI, 0.59-9.44) where an in-person assessment was judged necessary (eTable 28 in Supplement 1).

**Figure 4.**
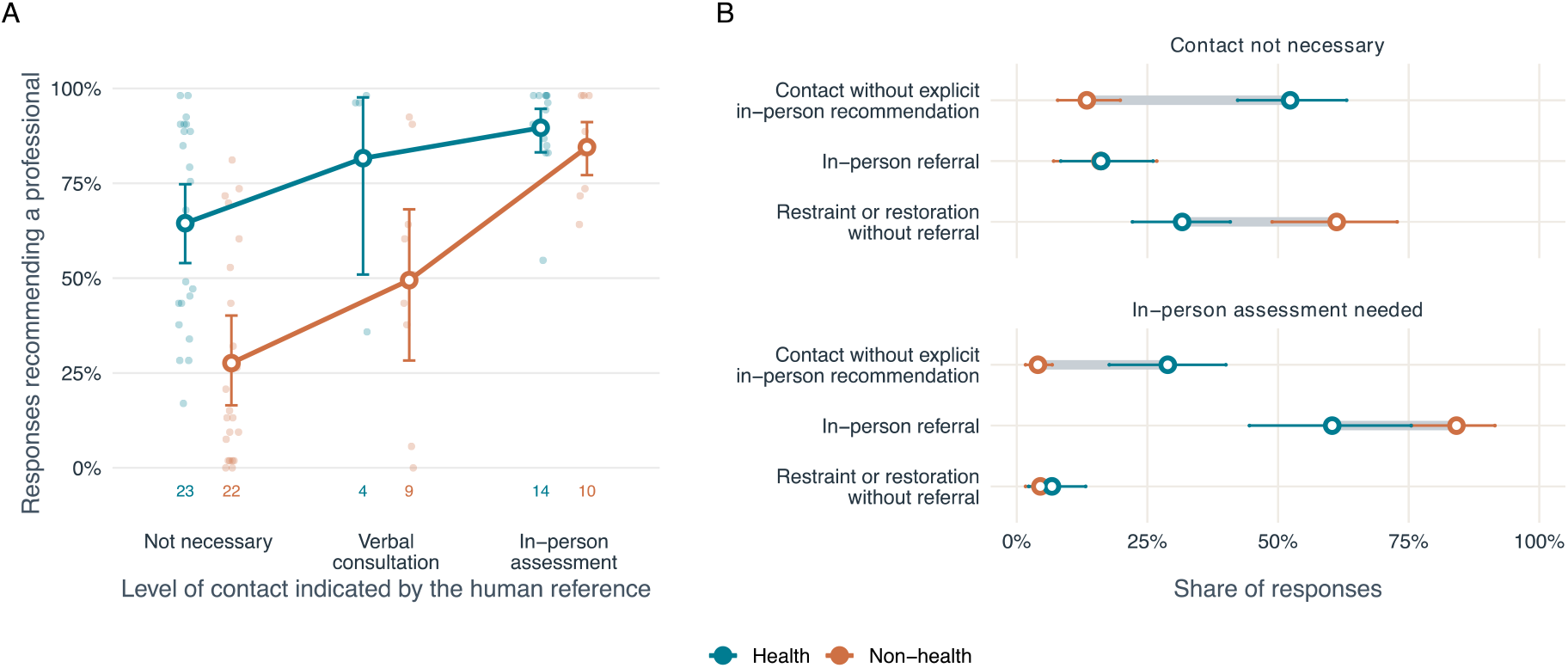
Professional Referral Against the Level of Contact Indicated by the Human Reference. A, Share of responses recommending a professional for each message (small points), by the level of professional contact the human reference judged necessary; large points are means across messages with 95% CIs, and the numbers below them are the messages at each level. B, Separately estimated recommendation rates at the 2 ends of the scale, where contact was judged not necessary and where an inperson assessment was judged necessary: contact without explicit in-person referral (remote or unspecified), in-person referral, and restraint or restoration of the reference practice without any referral. Points are means across messages with 95% CIs, and the gray bar joins the 2 sides; where the markers coincide (in-person referral where contact was not necessary, 16.2% and 16.0%), the health marker covers the non-health one. Medicine scenarios were assessed by 3 physicians and 2 pharmacists; other health scenarios by 3 physicians, with discussion of unresolved cases; the non-health reference is the consensus of 2 raters. Contact-mode rates average 2 coders and are not a partition of the 3-coder referral outcome. The 95% CIs are from 2,000 bootstrap resamples of messages.

Where an in-person assessment was judged necessary, referral was as frequent in health as outside it (89.6% and 84.5%). Health responses asked the person to be seen less often (60.4%; 95% CI, 44.5-75.5 vs 84.2%; 95% CI, 75.6-91.5; OR, 0.11; 95% CI, 0.01-0.89) and more often named a professional without asking for an assessment (28.9% vs 4.1%) (eTable 28 in Supplement 1).

Referral increased with the level of contact on both sides, less steeply in health (OR per level, 2.69; 95% CI, 1.47-4.91 vs 7.22; 95% CI, 3.70-14.07; likelihood-ratio P = .03; Spearman correlation across messages, 0.48 vs 0.65), so the association between need and referral was weaker in health. Unconditional referral rose with judged need on both sides (OR per level, 5.43; 95% CI, 2.80-10.51 vs 10.14; 95% CI, 4.78-21.50; Spearman correlation, 0.60 vs 0.71).

### 3.4 Release Date

The health-minus-non-health referral gap was present from the earliest configurations and grew until 2025: its mean was 20.5 percentage points among configurations released in 2023, 24.6 in 2024, 32.2 in 2025 and 32.3 in 2026 (eTable 8 in Supplement 1), a fitted increase of 4.8 percentage points per year (95% CI, 2.3-7.2) (Figure 5). Fitted referral rose in health from 68.6% to 79.1% and fell outside health from 49.9% to 43.7% (eTable 6 in Supplement 1). In health, fitted referral accompanied by intervention rose from 21.2% to 39.7% between the 2023 and 2026 ends of the comparison, and intervention without referral fell from 13.1% to 6.8%. The absence of a severity gradient in medicines and physical health persisted across release periods (correlations, −0.01, −0.02 and 0.09) (eTables 20 and 21 in Supplement 1).

**Figure 5.**
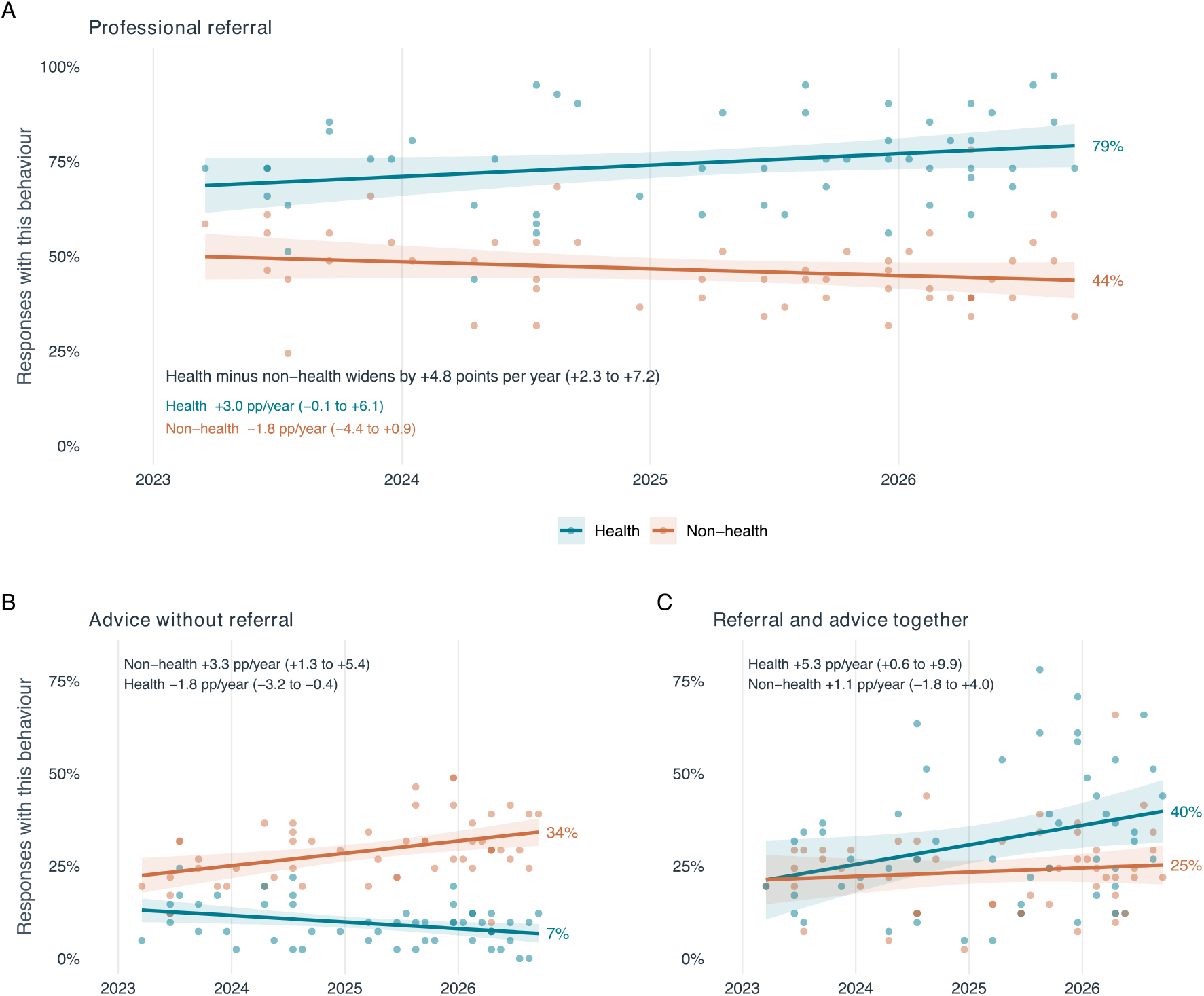
Professional Referral and Its Combination With Intervention by Release Date. Professional referral (A), self-directed intervention without any referral (B), and referral and intervention in the same response (C), against the release date of each configuration. Each point is one configuration on one side, with scenarios weighted equally; lines are least-squares fits on configuration rates and bands are 95% CIs; fitted values at the right-hand end are printed next to each line. Configurations are not independent draws, and access route is confounded with release date, so these trends describe an association with release date.

## 4 Discussion

Although no message asked whether to see a professional, every one of the 53 configurations recommended one more often for health than for comparable everyday problems. Against a human reference of the contact each situation required, the difference lay where no contact was needed, while where an assessment was needed the 2 domains referred at similar rates. Where physicians and pharmacists judged the instructions already received sufficient, 64.5% of health answers still told the person to consult a professional (46.3% only if things changed or the person was unsure), and 31.7% told the person to keep to the instructions; for comparable everyday problems the figures were 27.6% and 61.2%.

Where the person needed to be seen, about 9 health answers in 10 recommended a professional (89.6% vs 84.5%), and about 1 in 10 referred the person to no one; a consumer health tool built on a general-purpose model undertriaged 52% of emergencies (Ramaswamy et al. 2026). Health answers also asked for an in-person assessment less often than non-health answers (60.4% vs 84.2%), and in about 3 cases in 10 named a professional without saying whether a visit was needed, which leaves that decision to the person. With real requests one would also need to know what the person does after reading the answer. If people follow such advice, each unnecessary consultation could take an appointment in primary care, or a pharmacist’s time, from someone who needs it: risk-averse triage has added contacts wherever its effect on use has been measured (Boggan et al. 2020; Campbell et al. 2015), and use of these systems is higher among people without a usual source of primary care (Ayo-Ajibola et al. 2026), who are the least able to act on a generic instruction to ask their physician.

The health-minus-non-health difference in referral was already 20 percentage points in configurations released in 2023 and reached 32 points in those released in 2025 and 2026, as referral rose in health and fell outside it. Newer models also state their limitations less often (Sharma et al. 2025), and in our data they increasingly paired the referral in health with instructions on what to do in the meantime.

A general-purpose chatbot has no clinical history and cannot examine the person, and one built to triage would be a medical device requiring approval (Gilbert et al. 2023; Freyer et al. 2024). If the system tells a person that no care is needed and the situation then worsens, the provider may be held liable, which does not happen after a referral. For both reasons providers have an interest in keeping these systems cautious on health questions. Outside health, however, the systems could not see the boiler or the laptop either, and there their referral followed need. In health the same configurations attached a conditional referral to situations judged manageable more than twice as often (46.3% vs 21.0%), so the clause followed the topic of the question. A conditional referral is also what clinicians call safety netting, a recommended part of primary care consultations (Jones et al. 2019), but safety netting works when it names the signs to watch for and when to seek help (Smith et al. 2022), which a clause added by topic does not do. This design cannot say why. One possibility is that predicting the next token from training text is enough to produce the pattern, with no appraisal of the situation (Vannacci 2026a). The same consumer health tool overtriaged 64.8% of cases suitable for home care (Ramaswamy et al. 2026), and prompting models toward safety raised rated safety of advice and overtriage together (Xu et al. 2025).

Human triage also errs toward caution: of 5.3 million encounters in US emergency departments, 28.9% were overtriaged (Sax et al. 2023). The triage nurse, however, is responsible for that caution and can change the decision on seeing the patient. No one is responsible for the referral a chatbot adds, and nothing in the answer shows whether the situation was weighed; language models can likewise give moral verdicts that look right without following the reasons that would justify them (Vannacci et al. 2026).

Systems built for triage can be made to grade their advice. A self-triage system that guides a language model through clinical flowcharts followed the correct path in 99.1% of simulated conversations (Liu et al. 2026), and a model that retrieves triage guidelines and prior cases reduced overtriage from 28.8% to 12.7% of encounters against the consensus of triage nurses (Wong and Wong 2026). Both are given a triage task and a protocol to follow. A person who asks a general-purpose chatbot is alone with a system that has neither. Telling whether care is needed requires tools designed and validated for the purpose, and general-purpose chatbots were not built for it.

What people do with such advice is uncertain. In a randomized study of 1,298 members of the public, those assisted by a language model chose the right course of action in fewer than 44.2% of scenarios, no better than controls, although the models alone reached 56.3% (Bean et al. 2026). The advice also comes from systems that can confabulate, producing fluent guidance without a factual basis (Vannacci 2026b). When a patient arrives after such an exchange, what the system advised is part of the history, and patients who discuss online information with their clinician tend to report a better relationship (Tan and Goonawardene 2017); asking what the person was told to do before coming may be enough to bring it into the consultation.

### 4.1 Limitations

The messages were investigator-written and the configurations a selected panel; each configuration answered each message once, through an API or local deployment, under a brief English prompt. Outcomes and scenario ratings were coded by language models selected on agreement with expert human coding. The human reference was formed by a small panel, different in the 2 domains, that included the author of the messages; the main estimate was unchanged when single raters, or the external raters alone, replaced the consensus.

## 5 Conclusions

General-purpose language models, which are not built for triage, referred people to a professional when a visit was needed, in health as outside it, but in health also in 2 in 3 situations that physicians and pharmacists judged manageable without contact, mostly through a conditional clause used half as often outside health. Systems that people consult about whether to seek care should be evaluated on whether their advice distinguishes what can be deferred or managed at a distance, as well as on their referral of serious cases.

## Data Availability

The 82 messages, the 4,346 model responses with their coded labels, the scenario ratings, the human reference ratings and the analysis code in R and Python will be deposited in a public repository (Zenodo) with a permanent identifier at publication, with data under CC BY 4.0 and code under the MIT license (Supplement 2, Data Sharing Statement).

## Article Information

### Author Contributions

Dr Vannacci had full access to all of the data in the study and takes responsibility for the integrity of the data and the accuracy of the data analysis.

#### Concept and design

Vannacci, Ravaldi.

#### Acquisition, analysis, or interpretation of data

All authors.

#### Drafting of the manuscript

Vannacci.

#### Critical review of the manuscript for important intellectual content

All authors.

#### Statistical analysis

Vannacci.

### Conflict of Interest Disclosures

None reported. No author or author’s institution has received, in the past 36 months, payment or services from a third party that could be perceived to influence, or that give the appearance of potentially influencing, the submitted work.

### Funding/Support

This study received no specific funding. No author or author’s institution received payment or services from a third party for any aspect of the submitted work.

### Ethics

The study analyzed investigator-written messages and the answers of language models; it involved no patients, no human participants and no personal data, and did not require ethics committee review. It is not a clinical trial.

### Data Sharing Statement

See Supplement 2, in the supplementary material of this preprint.

### Additional Contributions

We thank Davide Arillotta, MD, Costanza Cacini, PharmD, and Andrea Madiai, Department of Neurosciences, Psychology, Drug Research and Child Health (NEUROFARBA), University of Florence, who rated the messages as external raters, the first 2 the health messages and the third the non-health messages. They received no compensation for their contribution.

## Declaration of generative AI and AI-assisted technologies in the manuscript preparation process

During the preparation of this work the authors used Claude Code (Anthropic) and ChatGPT Codex (OpenAI) to edit language and style, to search and check the literature and the accuracy of citations, and to write the analysis code and the code that draws the figures. The figures were drawn programmatically from the authors’ specifications; no generative AI image tools were used to create or alter them. The language models that were the object of the study, and those used as judges, are described in the Methods. After using these tools, the authors reviewed and edited the content as needed and take full responsibility for the content of the publication.

## Notes

### Competing Interest Statement

The authors have declared no competing interest.

